# Clustering of major depressive disorder genetic instruments identifies distinct and directionally opposing effects on cardiometabolic risk

**DOI:** 10.64898/2026.03.16.26347391

**Authors:** Dale Handley, Renu Bala, Francesco Casanova, Alexandra C Gillett, Chris WH Lo, Madhurbain Singh, Inês Barroso, Jack Bowden, Cathryn M Lewis, Jessica Tyrrell

**Author notes:** These authors contributed equally (Dale Handley, Renu Bala). Corresponding author: Dr. Dale Handley, Social, Genetic and Developmental Psychiatry Centre, Institute of Psychiatry, Psychology and Neuroscience, King’s College London, London.

## Abstract

**Background:** Major depressive disorder (MDD) is a highly heterogeneous condition that is frequently co-morbid with type 2 diabetes (T2D), and yet the biological mechanisms linking these diseases remain unclear. We aim to identify distinct biological pathways in depression that may modify T2D risk.

**Methods:** Using Clustered Mendelian randomisation (MR-Clust), we analysed 621 genome-wide significant MDD variants to identify clusters of variants with similar causal effects on T2D. These clusters were validated, and their causal effects were comprehensively tested against glycaemic traits, depression subtypes, T2D risk factors, and cardiometabolic biomarkers using external GWAS data and the UK Biobank. Functional annotation of these clusters was performed using FUMA.

**Results:** MR-Clust identified three distinct clusters of MDD-associated variants. Two clusters (MDD1 and MDD2) were causal for higher T2D and its related risk factors, adverse glycaemic and cardiometabolic profiles. Functional annotation implicated brain expression that overlapped strongly with depression-related traits such as smoking and neuroticism. By contrast, MDD3 was causal for lower T2D risk, more favourable glycaemic and cardiometabolic biomarker profiles, and was enriched for gene sets linked to fatty acid metabolism and steroid biosynthesis. MDD1 and MDD2 clusters were associated with atypical-like depression symptoms, whereas MDD3 was associated with melancholic depression symptoms.

**Conclusion:** Our findings demonstrate a heterogenous genetic architecture for depression, with distinct biological pathways conferring opposing effects on cardiometabolic health. Understanding this heterogeneity could help tailor prevention and treatment strategies for people with depression at greatest metabolic risk.

## Introduction

Depression is a highly heterogeneous condition. While many individuals experience only a single episode during their lifetime, others follow a recurrent course, and the most severe cases of major depressive disorder (MDD) are associated with a reduction in life expectancy of more than a decade (1). The Diagnostic and Statistical Manual of Mental Disorders 5 (DSM-5) lists nine symptoms for depression, with at least five required to meet criteria for MDD. This allows for over 250 unique symptom combinations (2), highlighting substantial clinical heterogeneity. MDD is a leading cause of disability worldwide and is associated with an increased lifetime risk of cardiometabolic diseases, including type 2 diabetes (T2D) (3,4). Given the relatively high prevalence of both MDD (∼11% lifetime occurrence) and T2D (∼9%), some co-occurrence would be expected; however, evidence suggests they co-occur at approximately twice the expected frequency (5–7). Several biological mechanisms may link MDD and T2D, including obesity, stress, chronic inflammation, and altered metabolic states. However, much of this evidence is derived from observational studies and is therefore vulnerable to confounding and reverse causality (8,9).

More recent studies, including our own, have applied Mendelian randomisation (MR) to investigate potential causal pathways between MDD and T2D (10,11). MR uses genetic variants as instrumental variables; because these variants are fixed at conception, bias due to confounding and reverse causation is reduced (12). Using MR, we previously demonstrated a clear causal bi-directional causal relationship between MDD and T2D (11). Notably, we observed clear heterogeneity in the causal effects of individual single nucleotide polymorphisms (SNPs). When analysing T2D as the exposure, grouping T2D-associated variants by biological pathway reduced heterogeneity and highlighted the importance of variants acting through obesity-mediated insulin resistance and adiposity (13).

There is also substantial genetic heterogeneity in SNP heritability across major depression subtypes, likely reflecting biological and etiological differences (14). A deeper understanding of this heterogeneity may provide insight into the pathophysiology of depression and clarify how depression influences downstream outcomes such as T2D. Given the phenotypic and genetic diversity of MDD, we hypothesise that MDD-associated variants act through distinct biological pathways, contributing to heterogeneity in their causal effects on T2D.

Clustered MR is a recently developed method that addresses heterogeneity in causal estimates by grouping variants with similar effects on an outcome (15). This approach is proposed to provide a more biologically informed perspective on causal associations (16). For example, clustered MR has shown that insulin-like growth factor 1 (IGF-1)-associated SNPs exhibit heterogeneous causal effects on T2D risk, likely due to differences in the molecular pathways underlying IGF-1 regulation (17).

Here, we build on our previous work by leveraging a recently published MDD GWAS including over 400,000 cases to: (a) replicate the causal association between MDD and T2D; (b) formally assess heterogeneity in this relationship; and (c) apply clustered MR to group MDD-associated variants according to their effects on T2D. We then replicate the identified clusters using external GWAS summary statistics and test their associations within the UK Biobank (UKB). Finally, we explore the biological pathways underlying these clustered causal effects using both summary statistics and the extensive phenotypic and genetic data available in UKB.

## Methods

### Clustered Mendelian randomisation

#### Summary statistics and genetic instruments

To instrument MDD genetically, we used genome-wide significant SNPs identified in 5,053,033 individuals of European ancestry from the largest available GWAS (18). In total, 621 independent autosomal variants were included as instruments for two-sample Mendelian randomisation (2SMR) (Supplementary Table 1) and extracted from the largest available T2D GWAS (n = 2,535,601) (13). Although there was minor sample overlap between these GWASs, we have previously shown this does not bias 2SMR causal estimates (11). Validation analyses used the FinnGen Release 12 “strict” T2D GWAS and an earlier DiaGRAM consortium T2D GWAS (19,20). A graphical overview of the study design is shown in Figure 1.

**Figure 1.**
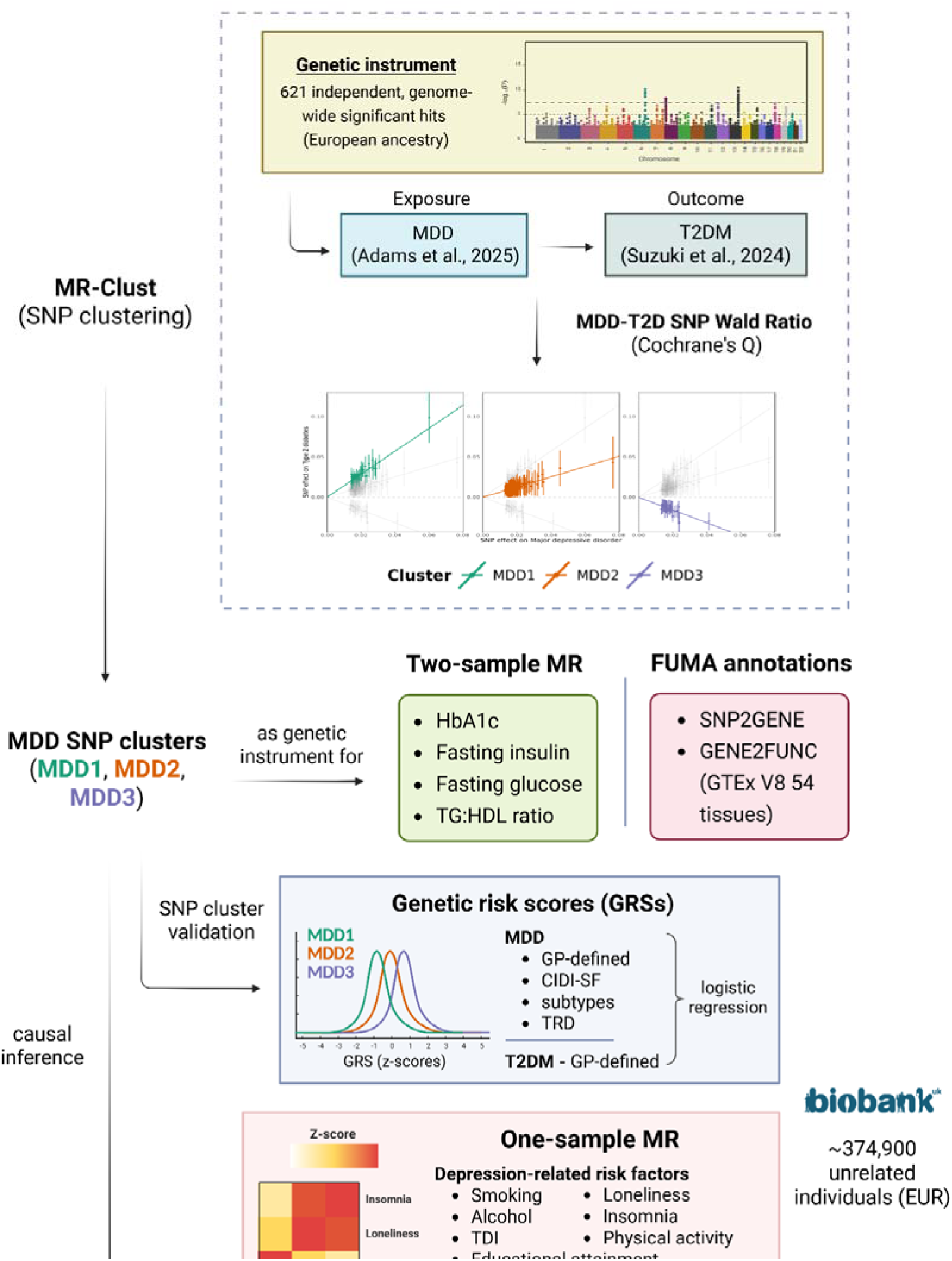
Graphical overview of the methods.

#### Statistical analyses

All MR analyses were conducted in R version 4.3.2. Effect sizes represent the unit change in the outcome per doubling in genetic liability to the exposure. P-values were adjusted for multiple comparisons using the Benjamini-Hochberg false discovery rate (FDR) method (5%), and all reported p-values are FDR-corrected.

We first performed two-sample Mendelian randomisation (2SMR) between MDD and T2D using the TwoSampleMR R package (21). Heterogeneity among SNP-specific Wald ratios was assessed using Cochran’s Q test, with p < 0.05 evidence of significant heterogeneity warranting further investigation.

Clustering of MDD-T2D SNPs was conducted using MR-Clust, which applies Gaussian mixture modelling to Wald ratio estimates to group variants with similar causal effects, potentially reflecting shared biological pathways (15). A posterior inclusion probability threshold of 0.8 and a minimum cluster size of five SNPs were required. Variants not meeting the probability threshold and clusters containing fewer than five SNPs were excluded. As a sensitivity analysis, clustering was repeated after removing SNPs failing Steiger filtering (22).

Clusters were validated using 2SMR in two stages. First, causal effects were re-estimated using the cluster discovery GWAS. Second, although no fully independent T2D GWAS of sufficient size without sample overlap was available, we used FinnGen and DIAGRAM T2D GWAS datasets to evaluate whether the identified MDD-T2D clusters were associated with T2D (19,20). MR-Egger regression was applied in all validation analyses to assess horizontal pleiotropy (23).

### Evaluating the causal relationship between MDD SNP clusters and measures of glycaemic control

#### Summary statistics and genetic instruments

The identified MDD-T2D cluster SNPs were extracted from GWAS summary statistics for key glycaemia-related traits, including glycated haemoglobin (HbA1c), fasting insulin, and fasting glucose in individuals with normoglycaemia (24,25). We also included triglyceride-to-high-density lipoprotein (HDL) ratio as a proxy for insulin resistance (26).

#### Statistical analysis

2SMR was performed to assess the causal effects of the identified MDD clusters on glycaemic traits. MR-Egger regression was used to evaluate horizontal pleiotropy.

### Function annotation of MDD-T2D clusters

We used FUMA (Functional Mapping and Annotation of Genome-Wide Association Studies) to functionally annotate each MDD-T2D cluster (27). SNPs were mapped to genes using the SNP2GENE function, and downstream analyses were conducted with GENE2FUNC. Specifically, we: (1) examined differential expression of cluster gene sets across 54 GTEx v8 tissues to assess tissue specificity; (2) tested for over-representation in pre-defined biological pathways from the Kyoto Encyclopedia of Genes and Genomes (KEGG) and Gene Ontology; and (3) identified overlap with gene sets reported in previously published GWASs from the GWAS Catalog (28).

### Validation and further interrogation of MDD clusters using MDD subtypes in the UKB

#### Cohort description

To validate our 2SMR findings and enable sex-stratified analyses, we conducted observational analyses of MDD and T2D risk in up to 374,900 unrelated individuals of European ancestry from the UK Biobank (UKB) (Supplementary Methods) (29). All analyses were adjusted for age at first assessment, sex, genotyping array, and the first ten principal components of ancestry. Sex-stratified analyses were also performed. Detailed trait definitions are provided in the Supplementary Methods.

#### Genetic risk score generation

To instrument MDD-T2D cluster-specific genetic liability, we generated weighted genetic risk scores (GRS) for each cluster separately using a publicly available pipeline implemented in PLINK v2.0 (30). Higher GRS values reflect greater genetic liability for the respective cluster.

#### Association of MDD-T2D clusters with MDD and T2D in the UKB

Logistic regression was used to test the association between the MDD-T2D cluster GRSs and both MDD and T2D, using two MDD definitions (GP-defined MDD, and Composite International Diagnostic Interview short form questionnaire (CIDI-SF)) (11).

#### Association of MDD-T2D clusters with depression subtypes in the UKB

To further understand the relationship between the MDD-T2D clusters and MDD, we tested the association of the three cluster GRSs with four depression subtypes,

– Melancholic depression, proxied by weight loss and insomnia during worst depressive episode (LWS, n = 5,424 controls and 4,482 cases);
– Atypical depression, proxied by weight gain and hypersomnia during the worst episode of depression (HWS, 9,109 controls and 797 cases);
– Treatment resistant depression according to the primary care electronic healthcare record and prescription data in individuals with GP-defined MDD (18,192 controls and 2,217 cases); and
– Lifetime severe depression according to the CIDI-SF (41,829 controls and 2,055 cases) (Supplementary methods).

### The causal relationship of MDD-T2D clusters with cardiometabolic risk factors in the UKB

To investigate mechanisms through which each cluster may influence T2D risk, we performed one-sample Mendelian randomisation (1SMR) to assess their causal effects on depression-related and cardiometabolic risk factors. Full phenotype definitions are provided in the Supplementary Methods.

### Depression-related risk factors

We analysed eleven risk factors: cigarettes smoked per day, ever smoking status, alcohol units per day, alcohol use disorder, Townsend deprivation index, educational attainment (years of education), loneliness, insomnia, and actigraphy-derived sedentary time, light-to-moderate physical activity, and moderate-to-vigorous physical activity.

#### Cardiometabolic risk factors

For cardiometabolic risk factors, we examined the causal effects of MDD-T2D cluster genetic liability on lipids (HDL, LDL, triglycerides, total cholesterol), adiposity (body mass index; BMI), glycaemic control (HbA1c), liver function (γ-glutamyl transferase; GGT, aspartate aminotransferase; AST, alanine aminotransferase; ALT), systemic inflammation (C-reactive protein; CRP), blood pressure (systolic; SBP and diastolic; DBP), and cardiometabolic diseases (coronary artery disease and stroke).

For body fat distribution traits, we assessed anthropometric measures (body fat percentage, waist circumference, hip circumference, waist-to-hip ratio, total adiposity), adipose tissue depot volumes (visceral adipose tissue; VAT, abdominal subcutaneous adipose tissue; aSAT, and the VAT:aSAT ratio), ectopic fat deposition and thigh muscle fat infiltration.

#### Statistical analysis

Since the MDD-T2D SNPs were generated using the MDD GWAS, the MDD-T2D GRSs were regressed onto the “GP-defined” MDD phenotype as the first stage of a two-stage least squares approach used for 1SMR. For continuous variables, the second stage was a linear regression, and for dichotomous variables, and logistic regression was used for the second stage of the two-stage least squares (31). To test whether the cluster-trait associations we observed were driven by differences in BMI, we used multivariable 1SMR (MVMR) to adjust for the genetic confounding effect of BMI for the cardiometabolic biomarkers and body fat distribution traits. A BMI GRS was generated using 73 BMI-associated SNPs from a BMI GWAS which has minimal sample overlap with the UKB and used to instrument BMI genetic liability (32).

## Results

A doubling in MDD genetic liability was strongly associated with increased T2D risk (OR: 1.17, 95% CI: 1.13-1.21, p = 4.7×10□¹□). Cochran’s Q indicated substantial heterogeneity among MDD-T2D SNP effects (Q = 4038.7, p < 1×10□²□□).

Given this heterogeneity, we applied MR-Clust to cluster variants with similar causal estimates. Three clusters were identified among SNPs associated with higher MDD genetic liability: MDD1 (very high T2D risk; 40 SNPs), MDD2 (moderately increased T2D risk; 123 SNPs), and MDD3 (reduced T2D risk; 28 SNPs) (Figure 2; Supplementary Table 2). Applying Steiger filtering prior to clustering removed the MDD1 cluster, resulting in a two-cluster solution with opposing directions of effect on T2D (Supplementary Figure 1).

**Figure 2.**
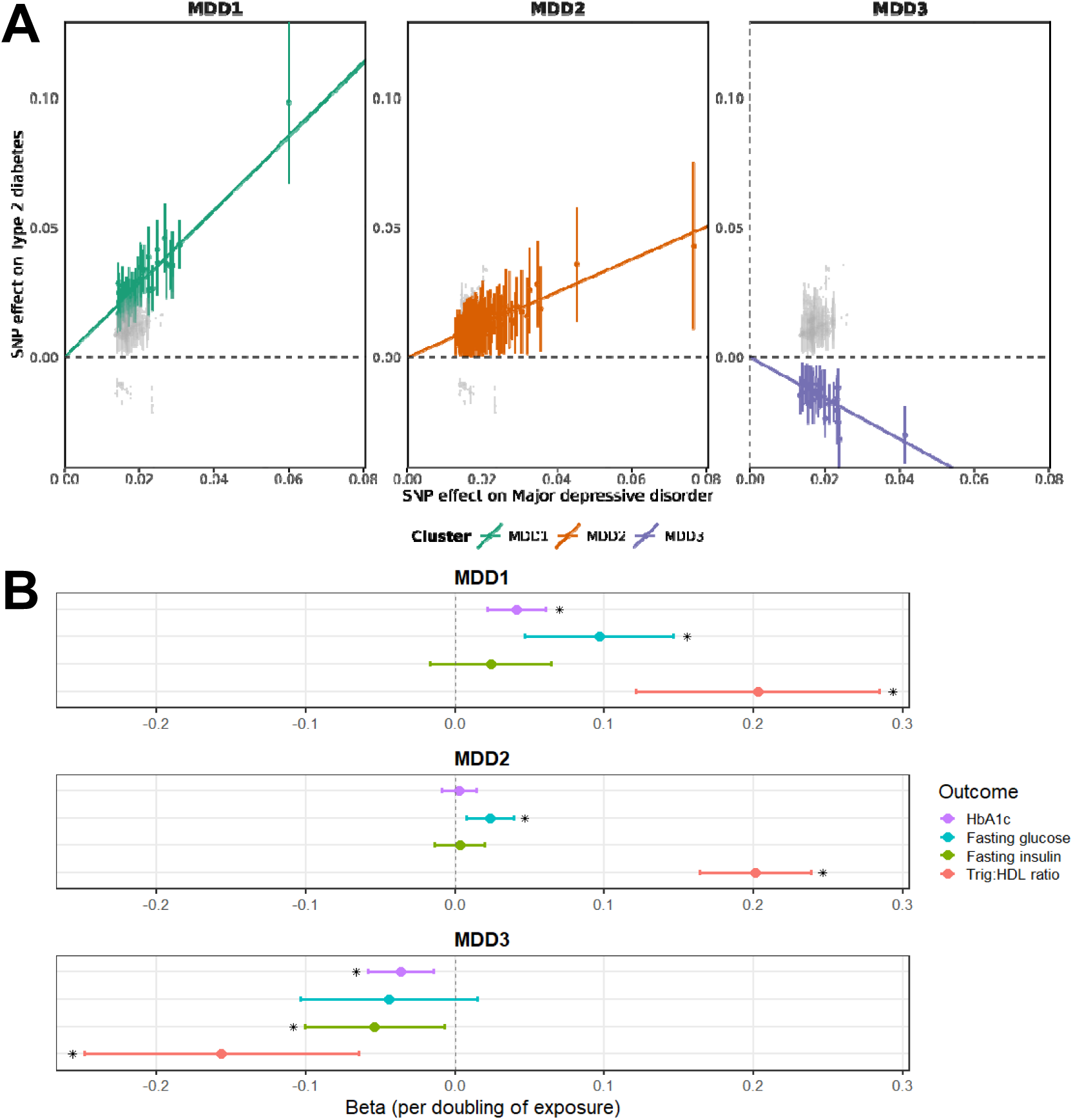
A) MDD-T2D SNP clusters identified using the MR-Clust approach and B) The causal effect of MDD-T2D clusters on glycaemic traits. SNP effect sizes are derived from GWASs for MDD and type 2 diabetes. Error bars represent standard errors reported in the MDD GWAS. Effect size represents beta per doubling of MDD genetic liability. Error bars represent 95% confidence intervals of the effect size. *, statistically significant (p < 0.05) after Benjamini-Hochberg FDR adjustment; Trig: HDL, Triglyceride to high density lipoprotein ratio.

We replicated these findings using 2SMR in the discovery T2D GWAS and two additional T2D GWAS datasets (13,19,20). Higher MDD1 genetic liability was strongly associated with increased T2D risk in the discovery GWAS (OR: 2.68, 95% CI: 2.54-2.83, p = 2.9×10□²□²), FinnGen (Release 12) (OR: 2.22, 95% CI: 1.96-2.53, p = 7.9×10□³□), and DiaGRAM (OR: 2.71, 95% CI: 2.40-3.07, p = 2.0×10□□□). Similarly, MDD2 liability increased T2D risk, whereas MDD3 liability decreased T2D risk across all datasets (Supplementary Figure 2). MR-Egger provided no evidence of horizontal pleiotropy (Supplementary Table 3).

### MDD-T2D SNP clusters show distinct causal profiles across glycaemic traits

Given the opposing causal effects of these SNP clusters on T2D risk, we examined whether similar patterns were observed for glycaemic traits in normoglycaemic individuals using MAGIC consortium summary statistics. Higher MDD1 genetic liability was associated with increased HbA1c (β = 0.044, SE = 0.012, p = 8.4×10□□), fasting glucose (β = 0.097, SE = 0.025, p = 1.4×10□□), and triglyceride-to-HDL ratio (β = 0.097, SE = 0.025, p = 1.4×10□□).

Higher MDD2 liability increased fasting glucose (β = 0.023, SE = 0.008, p = 8.3×10□³) and triglyceride-to-HDL ratio (β = 0.021, SE = 0.019, p = 1.6×10□²□) but was not associated with HbA1c (β = 0.002, SE = 0.006, p = 0.71). In contrast, higher MDD3 liability was associated with lower HbA1c (β = –0.036, SE = 0.011, p = 2.3×10□³), fasting insulin (β = –0.054, SE = 0.024, p = 0.031), and triglyceride-to-HDL ratio (β = –0.046, SE = 0.038, p = 0.031) (Figure 2). MR-Egger indicated evidence of horizontal pleiotropy only for the association between MDD2 and fasting glucose (p = 1.4×10□³) (Supplementary Table 4).

### MDD-T2D SNP clusters show different tissue-specific expression and show distinct gene set overlap across a variety of pre-defined traits

Given the distinct causal effects of the MDD-T2D clusters on T2D risk and glycaemic traits, we investigated whether they differed in tissue expression and biological pathway enrichment using FUMA.

The MDD1 gene set was overexpressed in the cerebellar hemisphere (p = 8.2×10□□) and overlapped with depression-related traits, including neuroticism (p = 2.1×10□¹³) and alcohol use disorder (p = 4.7×10□□) (Supplementary Figures 3 and 4). Similarly, MDD2 genes showed increased expression across several brain regions and overlapped with depression-related phenotypes, including adult body size (p = 8.6×10□□), smoking status (p = 1.9×10□□), and chronotype (p = 3.2×10□□) (Supplementary Figures 5 and 6). No pathway enrichment was identified for MDD1 or MDD2.

**Figure 4.**
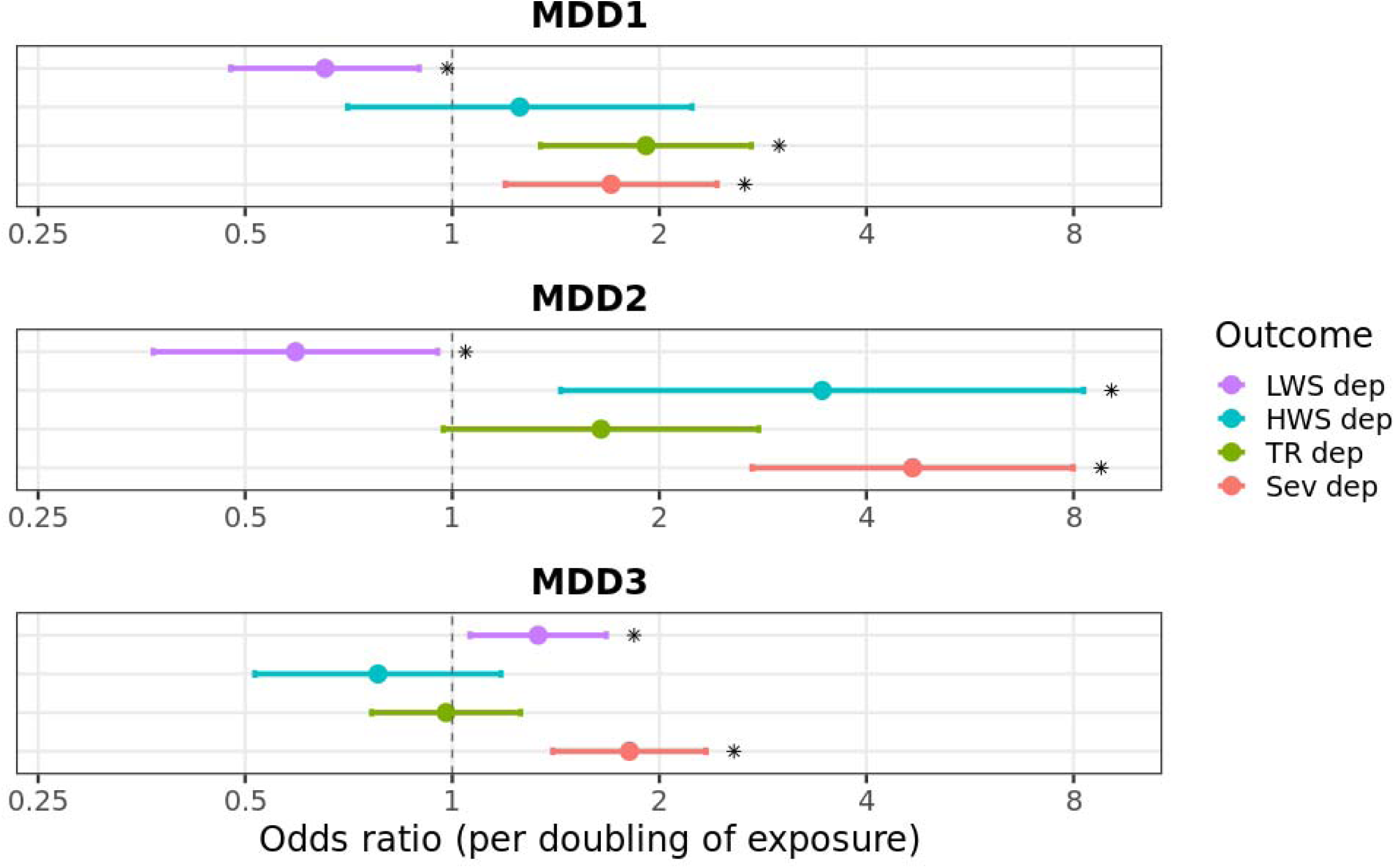
The association between MDD-T2D SNP clusters and depression subtypes. Effect sizes represent odds ratio per doubling in MDD genetic liability. Error bars represent 95% confidence intervals of the estimate. *, statistically significant after Benjamini-Hochberg FDR adjustment (p < 0.05). LWS dep, low weight and sleep depression; HWS dep, High weight and sleep depression; TR dep, Treatment resistant depression; Sev dep, ever severely depressed.

**Figure 5.**
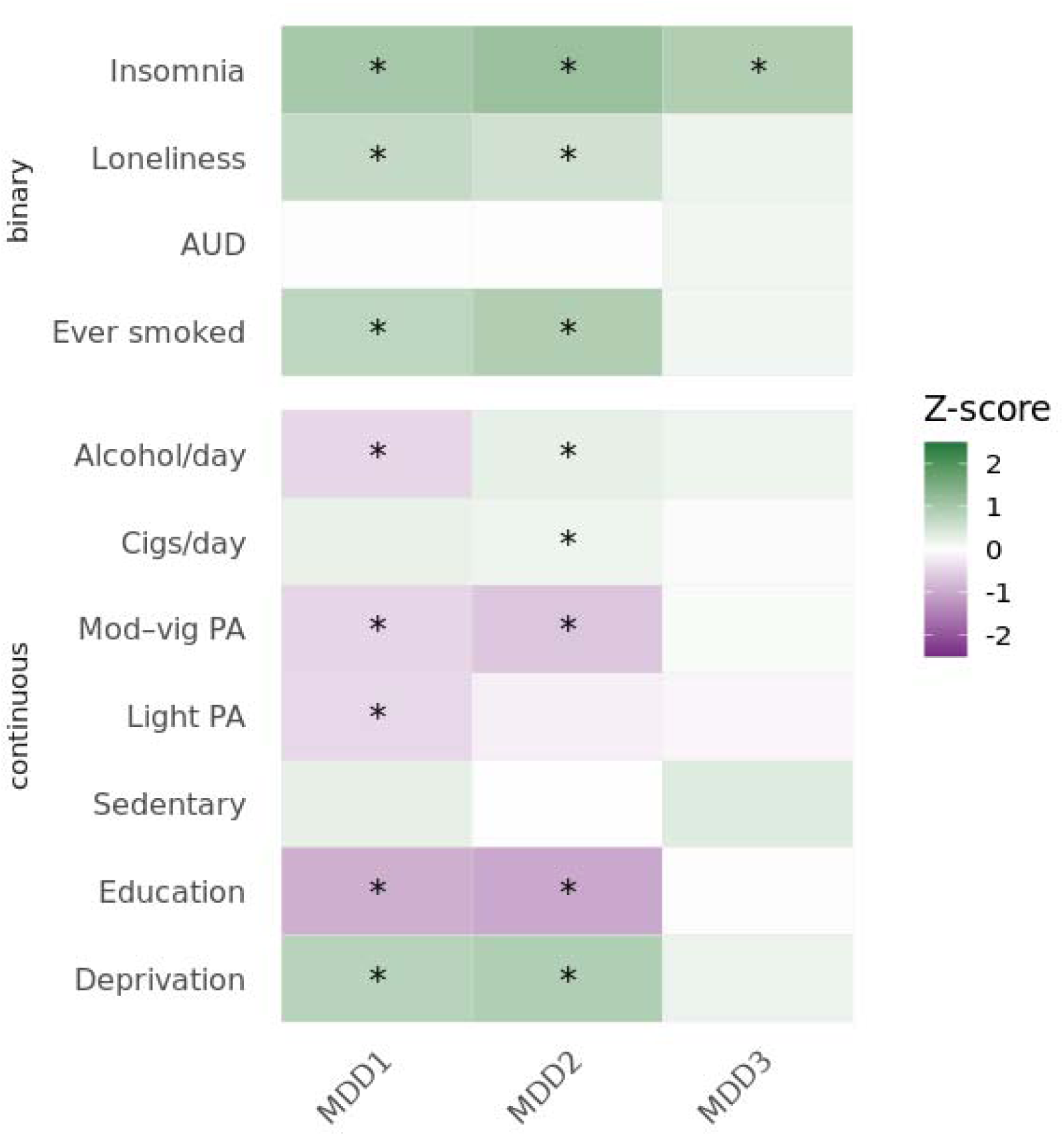
Relationship between health and lifestyle factors and the causal effects of MDD-T2D SNP clusters. Effect sizes are presented as Z scores standardised for the number of variants in each cluster. *, statistically significant after Benjamini-Hochberg FDR adjustment (p < 0.05). AUD, alcohol use disorder; Cigs, cigarettes; Mod-vig PA, Moderate to vigorous physical activity; Light PA, light physical activity; Sedentary, sedentary time; Education, education years; Deprivation, Townsend Deprivation Index.

**Figure 6.**
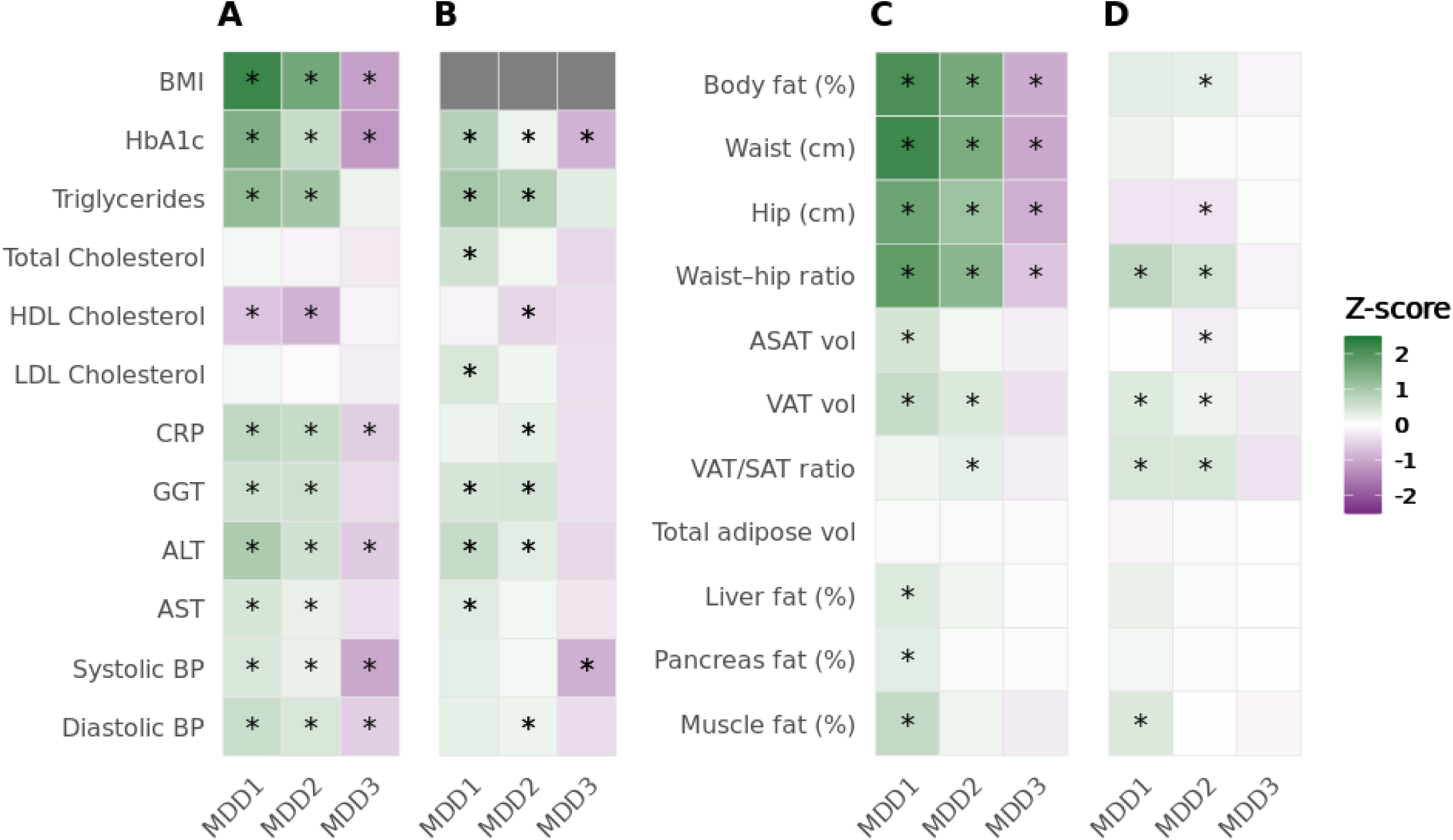
Heatmap of the causal effects of MDD-T2D SNP clusters on cardiometabolic biomarkers. A) unadjusted for BMI and B) adjusted for BMI genetic liability and on different adiposity measures C) unadjusted for BMI and D) adjusted for BMI genetic liability. For easier comparison, the effect sizes are presented as Z scores standardised for the number of variants in each cluster. *, statistically significant after Benjamini-Hochberg FDR adjustment (p < 0.05). BMI, body mass index; HbA1c, glycated haemoglobin; CRP, C-reactive protein; GGT, γ-glutamyl transferase; ALT, alanine aminotransferase; AST, aspartate transaminase; BP, blood pressure. Waist, waist circumference; hip, hip circumference; ASAT, abdominal subcutaneous adipose tissue; VAT, visceral adipose tissue; liver fat, percentage liver proton density fat fraction; pancreas fat, percentage pancreas proton density fat fraction; muscle fat, muscle fat infiltration; vol, volume

MDD3 demonstrated a distinct expression profile. Although no tissues survived multiple testing correction, MDD3 genes were nominally overexpressed in the adrenal gland (p = 0.011) and under-expressed in skeletal muscle (p = 9.7×10□³). The MDD3 gene set showed strong enrichment for plasma omega-3 polyunsaturated fatty acids (p = 1.6×10□□), omega-6 polyunsaturated fatty acids (p = 3.8×10□□), and hypertriglyceridaemia (p = 2.2×10□□), and was enriched in steroid and cholesterol metabolism pathways in both KEGG and Gene Ontology (Supplementary Figures 7-9).

### MDD-T2D SNP clusters validate in the UKB and are associated with different depression subtypes

To further validate our 2SMR findings, we assessed associations between the MDD-T2D SNP clusters and alternative definitions of MDD and T2D in UKB. As expected, higher MDD1 and MDD2 genetic liability were associated with increased risk of both MDD (MDD1 OR: 1.87, 95% CI: 1.58-2.20, p = 2.0×10□¹³; MDD2 OR: 3.82, 95% CI: 2.98-4.92, p = 1.42×10□²□□) and T2D (MDD1 OR: 2.11, 95% CI: 1.85-2.42, p = 1.8×10□²□□; MDD2 OR: 3.24, 95% CI: 2.64-3.97, p = 6.99×10□³□□). In contrast, higher MDD3 liability was associated with increased MDD risk (OR: 1.30, 95% CI: 1.13-1.51, p = 4.09×10□□) but reduced T2D risk (OR: 0.86, 95% CI: 0.76-0.97, p = 0.012). No notable sex differences were observed (Supplementary Figure 9).

We next examined associations with depression subtypes to explore potential explanations for the observed metabolic differences. All clusters were associated with increased risk of severe depression (p < 0.05 for all). The atypical depression phenotype characterised by low weight and sleep was associated with lower risk in MDD1 and MDD2 (MDD1 OR: 0.65, 95% CI: 0.47-0.89, p = 8.1×10□³; MDD2 OR: 0.59, 95% CI: 0.37-0.95, p = 0.031) and higher risk in MDD3 (OR: 1.33, 95% CI: 1.06-1.67, p = 0.013) (Figure 3). These patterns were largely consistent across sexes (Supplementary Figures 10 and 11).

### MDD-T2D SNP clusters are causal for depression-related cardiometabolic risk factors

Given the associations with distinct depression subtypes, we next examined whether the clusters were linked to established depression-related cardiometabolic risk factors. Higher MDD1 and MDD2 genetic liability were associated with increased risk of eight of the eleven risk factors examined, including insomnia, fewer years of education, ever smoking, and greater deprivation (adjusted p < 0.05 for all) (Figure 4; Supplementary Table 5). In contrast, MDD3 liability was associated only with increased risk of insomnia (OR: 1.20, 95% CI: 1.11-1.29, p = 2.1×10□□). Sex-stratified analyses revealed no clear differences (Supplementary Figure 12).

### MDD-T2D SNP clusters are causal for different cardiometabolic profiles independent of BMI

We next examined the effects of genetically predicted MDD-T2D cluster liability on cardiometabolic risk factors. MR analyses indicated that higher MDD1 and MDD2 liability were associated with poorer cardiometabolic profiles, reflected in adverse biomarker patterns (Figure 5a).

Higher MDD1 liability was associated with increased BMI (β = 2.55, SE = 0.18, p = 4.1×10□□□), triglycerides (β = 0.33, SE = 0.07, p = 1.2×10□¹□), CRP (β = 0.77, SE = 0.17, p = 6.3×10□□), SBP (β = 1.00, SE = 0.37, p = 6.6×10□³), and DBP (β = 1.08, SE = 0.21, p = 3.7×10□□), and with lower HDL (β = –0.06, SE = 0.014, p = 2.0×10□□). A similar biomarker profile was observed for MDD2 (Figure 5b). Conversely, higher MDD3 liability showed a consistent trend toward a more favourable metabolic profile, with lower BMI (β = –2.16, SE = 0.44, p = 6.8×10□□), CRP (β = –0.73, SE = 0.24, p = 2.5×10□³), SBP (β = –5.18, SE = 0.98, p = 1.2×10□□), and DBP (β = –1.67, SE = 0.56, p = 2.9×10□³) (Supplementary Table 6).

After BMI adjustment, higher MDD1 liability remained associated with increased HbA1c (β = 1.50, SE = 0.28, p = 8.6×10□□), triglycerides (β = 0.28, SE = 0.04, p = 1.0×10□¹□), and liver enzymes (p < 0.05). MDD2 liability was newly associated with higher total cholesterol (β = 0.17, SE = 0.049, p = 5.3×10□□) and LDL cholesterol (β = 0.11, SE = 0.038, p = 3.9×10□³), and remained associated with higher HbA1c (β = 0.36, SE = 0.15, p = 0.18), triglycerides (β = 0.22, SE = 0.023, p = 3.0×10□²¹), CRP (β = 0.31, SE = 0.11, p = 2.44×10□³), liver enzymes (p < 0.05 for all), DBP (β = 0.54, SE = 0.24, p = 0.022), and lower HDL (β = –0.045, SE = 0.008, p = 1.2×10□□).

Higher MDD3 liability remained associated with lower HbA1c (β = –1.58, SE = 0.34, p = 4.4×10□□) and SBP (β = –4.53, SE = 0.94, p = 1.47×10□□).

MDD1 and MDD2 were also associated with increased risk of stroke and coronary artery disease (p < 0.05), whereas MDD3 was not (p > 0.05); these effects persisted also after BMI adjustment (Supplementary Table 7; Supplementary Figure 13).

### MDD-T2D SNP clusters are causal for differences in body fat distribution independent of BMI

Before BMI adjustment, higher MDD1 and MDD2 were associated with unfavourable fat distribution. Higher liability to both clusters was associated with increased body fat percentage, waist-to-hip ratio, and VAT volume (p < 0.05 for all). Higher MDD1 genetic liability was additionally associated with greater hepatic (β = 2.38, SE = 1.16, p = 0.038) and pancreatic fat deposition (β = 1.75, SE = 0.70, p = 0.013), as well as increased thigh muscle fat infiltration (β = 0.94, SE = 0.22, p = 2.1×10□□). Higher MDD2 genetic liability was associated with a higher VAT:aSAT ratio (β = 0.049, SE = 0.016, p = 1.7×10□³). Higher MDD3 liability was associated with a more favourable fat distribution, including lower body fat percentage (β = –1.76, SE = 0.34, p = 2.61×10□□) and waist-to-hip ratio (β = –0.013, SE = 0.004, p = 3.82×10□□), but not with other adiposity traits (Figure 5c; Supplementary Table 8). After adjusting for BMI genetic liability, MDD1 and MDD2 remained associated with unfavourable fat distribution (Figure 5d; Supplementary Table 9; Supplementary Figure 14).

## Discussion

Applying clustered MR to publicly available MDD and T2D summary statistics, we identified three distinct clusters of depression-associated variants with divergent causal effects on T2D risk. MDD1 and MDD2 were associated with increased T2D risk, whereas MDD3 was associated with reduced risk. These findings were replicated across multiple GWAS datasets and in UKB, and were supported by consistent differences in glycaemic traits, depression-related cardiometabolic risk factors, biomarker profiles, and body fat distribution. Collectively, our results indicate that the relationship between depression and cardiometabolic risk is not uniform, but instead reflects multiple, and in some cases opposing biological pathways.

The MDD1 and MDD2 clusters were enriched for loci expressed in the brain and showed strong overlap with behavioural and affective traits, including smoking, neuroticism, and alcohol use. These patterns suggest that these clusters capture neurobiological pathways related to emotional regulation, reward processing, and stress reactivity, which are core mechanisms disrupted in depression (33).Their association with lower risk of typical (low WS) depressive subtype may indicate that they reflect an atypical “immunometabolic” depression phenotype, clinically characterised by hyperphagia, hypersomnia, and frequent co-occurrence with obesity, low-grade chronic inflammation, and metabolic dysregulation (34). These clusters may also capture behavioural pathways linked to self-medication in depression, which further compound metabolic risk. Nicotine and alcohol influence appetite regulation, hepatic metabolism, and glucose homeostasis, potentially exacerbating underlying genetic susceptibility to metabolic dysfunction (35–37).Together, both central and peripheral mechanisms likely contribute to the cardiometabolic impairment associated with increased genetic liability in these clusters.

MDD3 exhibited a distinct biological and phenotypic profile. This cluster was associated with reduced T2D risk, favourable glycaemic and lipid profiles, and lower systemic inflammation and adiposity. It was also linked to increased risk of melancholic (low WS) depression, which is generally less metabolically disruptive than atypical depression (38,39). These findings are consistent with evidence that depression subtypes have distinct genetic architectures, suggesting that MDD3 may reflect a genetic loading toward less metabolically adverse depressive features.

MDD3 showed nominal overexpression in the adrenal gland and was enriched in pathways involved in cholesterol and steroid hormone pathways and had strong overlap with plasma polyunsaturated fatty acid (PUFA) levels. PUFAs influence neuronal membrane fluidity and synaptic transmission, while steroid metabolism plays a central role in systemic metabolic regulation (40–42). Increasing evidence implicates PUFA biology in MDD, with lower omega-3 levels, higher omega-6 levels, and impaired PUFA metabolism being associated with increased MDD risk, insulin resistance, and adiposity (43). MDD3’s association with higher omega-3 derived fatty acids may therefore link improved emotional regulation with enhanced insulin sensitivity. MDD3 remained associated with lower HbA1c and blood pressure after BMI adjustment, suggesting that its protective effects reflect intrinsic metabolic regulation rather than differences in adiposity.

Individuals with higher MDD1 or MDD2 genetic liability may represent a subgroup with vulnerability to depression accompanied by obesity, insulin resistance, and broader cardiometabolic dysfunction. These individuals may benefit from proactive cardiometabolic monitoring and targeted weight management, particularly when obesity is present or emerging. In opposition to this, individuals with higher MDD3 liability may derive greater benefit from interventions focused on emotional regulation and neuroendocrine balance rather than metabolic control. These findings highlight the potential value of stratifying patients by genetic and biological profiles when designing prevention and treatment strategies for depression and cardiometabolic disease.

A key strength of this study is the use of large, publicly available datasets alongside extensive validation and sensitivity analyses, enhancing the robustness of the identified clusters. We further characterised cluster effects across a broad range of phenotypes related to both MDD and T2D, integrated with functional annotation, thus enabling mechanistic inference through converging lines of evidence.

Several limitations warrant consideration. First, although MR-Clust is a well-established method and we conducted multiple sensitivity analyses, horizontal pleiotropy cannot be fully excluded. Second, our analyses were restricted to individuals of European ancestry, limiting generalisability across populations. Third, depression subtypes derived from UKB mental health questionnaire data rely on retrospective self-report and may be subject to misclassification. Finally, while functional annotation provides biological insight, experimental validation is required to confirm the underlying mechanisms of these clusters.

This study demonstrates that the causal relationship between depression and T2D is not uniform, but instead reflects multiple, interconnected, and at times opposing biological pathways. Using clustered MR, we identified three biologically distinct groups of depression-associated variants: two clusters (MDD1 and MDD2) that increase T2D risk through neurobehavioural and immunometabolic mechanisms, and a third cluster (MDD3) is metabolically protective and enriched for pathways related to fatty acid metabolism and steroid biosynthesis. These findings show that the metabolic consequences of depression depend strongly on its underlying biological architecture, helping to explain why some individuals with depression develop metabolic disease while others do not. Realising this heterogeneity opens opportunities for more targeted prevention and treatment strategies, including prioritising cardiometabolic monitoring in individuals with high MDD1/MDD2 genetic liability.

## Acknowledgements and funding

This work was funded by MRC project grant MR/X009815/1. This study was supported by the National Institute for Health and Care Research Exeter Biomedical Research Centre and by the NIHR Maudsley Biomedical Research Centre at South London and Maudsley NHS Foundation Trust and King’s College London. The views expressed are those of the author(s) and not necessarily those of the NIHR or the Department of Health and Social Care.

## Manuscript contributions

DH contributed to the conception of the study, performed the analysis, and prepared and revised the manuscript. RB contributed to the study conception and primary data analysis, aided in manuscript preparation, and revised the manuscript. FC contributed to the study conception and revised the manuscript. ACG aided in acquiring project funding, contributed to study conception and revised the manuscript. CWHL contributed to study conception, data management, phenotype definitions, and revised the manuscript. MS aided in primary analysis and interpretation and manuscript revision. IB aided in conceiving the study and revised the manuscript. JB aided in acquiring funding and revised the manuscript. CML aided in acquiring funding and developing the study direction, project management, and revised the manuscript. JT contributed to conception of the study, aided in funding acquisition, aided in leading the study direction, and prepared and revised the manuscript.

## Ethics statement

This research was conducted using the UK Biobank resource under approved application 103356. UK Biobank has received ethical approval from the North West Multi-centre Research Ethics Committee (MREC) (reference 11/NW/0382), and all participants provided written informed consent prior to participation. The present analyses were conducted in accordance with the principles of the Declaration of Helsinki.

## Data availability

All summary statistics from GWAS described in this manuscript are publicly available. Access to UK biobank data is made available following successful application.

## Previous data presentations

This data has not been presented or disseminated outside of a centre talk at the author’s home institution.

## References

1. Laursen TM, Musliner KL, Benros ME, Vestergaard M, Munk-Olsen T. Mortality and life expectancy in persons with severe unipolar depression. J Affect Disord. 2016 Mar 15;193:203–7.

2. American Psychiatric Association. Diagnostic and Statistical Manual of Mental Disorders (DSM-5®). American Psychiatric Publishing; 2013.

3. Zeng J, Qiu Y, Yang C, Fan X, Zhou X, Zhang C, et al. Cardiovascular diseases and depression: A meta-analysis and Mendelian randomisation analysis. Mol Psychiatry. 2025 Sep;30(9):4234–46.

4. Mezuk B, Eaton WW, Albrecht S, Golden SH. Depression and type 2 diabetes over the lifespan: a meta-analysis. Diabetes Care. 2008 Dec;31(12):2383–90.

5. Lim R, Beekley A, Johnson DC, Davis KA. Early and late complications of bariatric operation. Trauma Surg Acute Care Open. 2018 Oct 1;3(1):e000219.

6. Zheng Y, Ley SH, Hu FB. Global aetiology and epidemiology of type 2 diabetes mellitus and its complications. Nat Rev Endocrinol. 2018 Feb;14(2):88–98.

7. Holt RIG, de Groot M, Golden SH. Diabetes and Depression. Curr Diab Rep. 2014 Apr 18;14(6):491.

8. Moulton CD, Pickup JC, Ismail K. The link between depression and diabetes: the search for shared mechanisms. Lancet Diabetes Endocrinol. 2015 Jun 1;3(6):461–71.

9. Beverly EA, Gonzalez JS. The Interconnected Complexity of Diabetes and Depression. Diabetes Spectr. 2025 Feb 14;38(1):23–31.

10. Zhang M, Chen J, Yin Z, Wang L, Peng L. The association between depression and metabolic syndrome and its components: a bidirectional two-sample Mendelian randomisation study. Transl Psychiatry. 2021 Dec 13;11(1):633.

11. Bala R, Handley D, Gillett A, Green H, Bowden J, Wood A, et al. Evidence of bidirectional relationship between type 2 diabetes and depression; a Mendelian randomisation study. Mol Psychiatry. 2025 Jul 1;1–11.

12. Sanderson E, Glymour MM, Holmes MV, Kang H, Morrison J, Munafò MR, et al. Mendelian randomisation. Nat Rev Methods Primer. 2022 Feb 10;2(1):6.

13. Suzuki K, Hatzikotoulas K, Southam L, Taylor HJ, Yin X, Lorenz KM, et al. Genetic drivers of heterogeneity in type 2 diabetes pathophysiology. Nature. 2024 Mar;627(8003):347–57.

14. Nguyen TD, Harder A, Xiong Y, Kowalec K, Hägg S, Cai N, et al. Genetic heterogeneity and subtypes of major depression. Mol Psychiatry. 2022 Mar;27(3):1667–75.

15. Foley CN, Mason AM, Kirk PDW, Burgess S. MR-Clust: clustering of genetic variants in Mendelian randomisation with similar causal estimates. Bioinformatics. 2021 May 1;37(4):531–41.

16. Verkouter I, de Mutsert R, Smit RAJ, Trompet S, Rosendaal FR, van Heemst D, et al. The contribution of tissue-grouped BMI-associated gene sets to cardiometabolic-disease risk: a Mendelian randomisation study. Int J Epidemiol. 2020 Aug 1;49(4):1246–56.

17. Wang W, Tesfay EB, van Klinken JB, Willems van Dijk K, Bartke A, van Heemst D, et al. Clustered Mendelian randomisation analyses identify distinct and opposing pathways in the association between genetically influenced insulin-like growth factor-1 and type 2 diabetes mellitus. Int J Epidemiol. 2022 Jun 3;51(6):1874–85.

18. Adams MJ, Streit F, Meng X, Awasthi S, Adey BN, Choi KW, et al. Trans-ancestry genome-wide study of depression identifies 697 associations implicating cell types and pharmacotherapies. Cell. 2025 Feb 6;188(3):640–652.e9.

19. Kurki MI, Karjalainen J, Palta P, Sipilä TP, Kristiansson K, Donner KM, et al. FinnGen provides genetic insights from a well-phenotyped isolated population. Nature. 2023 Jan;613(7944):508–18.

20. Mahajan A, Taliun D, Thurner M, Robertson NR, Torres JM, Rayner NW, et al. Fine-mapping type 2 diabetes loci to single-variant resolution using high-density imputation and islet-specific epigenome maps. Nat Genet. 2018 Nov;50(11):1505–13.

21. Hemani G, Zheng J, Elsworth B, Wade KH, Haberland V, Baird D, et al. The MR-Base platform supports systematic causal inference across the human phenome. Loos R, editor. eLife. 2018 May 30;7:e34408.

22. Hemani G, Tilling K, Davey Smith G. Orienting the causal relationship between imprecisely measured traits using GWAS summary data. PLoS Genet. 2017 Nov 17;13(11):e1007081.

23. Burgess S, Thompson SG. Interpreting findings from Mendelian randomisation using the MR-Egger method. Eur J Epidemiol. 2017;32(5):377–89.

24. Scott RA, Lagou V, Welch RP, Wheeler E, Montasser ME, Luan J, et al. Large-scale association analyses identify new loci influencing glycemic traits and provide insight into the underlying biological pathways. Nat Genet. 2012 Sep;44(9):991–1005.

25. Wheeler E, Leong A, Liu CT, Hivert MF, Strawbridge RJ, Podmore C, et al. Impact of common genetic determinants of Hemoglobin A1c on type 2 diabetes risk and diagnosis in ancestrally diverse populations: A transethnic genome-wide meta-analysis. PLOS Med. 2017 Sep 12;14(9):e1002383.

26. Oliveri A, Rebernick RJ, Kuppa A, Pant A, Chen Y, Du X, et al. Comprehensive genetic study of the insulin resistance marker TG:HDL-C in the UK Biobank. Nat Genet. 2024 Feb;56(2):212–21.

27. Watanabe K, Taskesen E, van Bochoven A, Posthuma D. Functional mapping and annotation of genetic associations with FUMA. Nat Commun. 2017 Nov 28;8(1):1826.

28. Sollis E, Mosaku A, Abid A, Buniello A, Cerezo M, Gil L, et al. The NHGRI-EBI GWAS Catalog: knowledgebase and deposition resource. Nucleic Acids Res. 2022 Nov 9;51(D1):D977–85.

29. Bycroft C, Freeman C, Petkova D, Band G, Elliott LT, Sharp K, et al. The UK Biobank resource with deep phenotyping and genomic data. Nature. 2018 Oct;562(7726):203–9.

30. Chang CC, Chow CC, Tellier LC, Vattikuti S, Purcell SM, Lee JJ. Second-generation PLINK: rising to the challenge of larger and richer datasets. GigaScience. 2015;4:7.

31. Sanderson E, Davey Smith G, Windmeijer F, Bowden J. An examination of multivariable Mendelian randomisation in the single-sample and two-sample summary data settings. Int J Epidemiol. 2018 Dec 10;48(3):713–27.

33. Dean J, Keshavan M. The neurobiology of depression: An integrated view. Asian J Psychiatry. 2017 Jun 1;27:101–11.

34. Penninx BWJH, Lamers F, Jansen R, Berk M, Khandaker GM, Picker LD, et al. Immuno-metabolic depression: from concept to implementation. Lancet Reg Health – Eur [Internet]. 2025 Jan 1 [cited 2025 Nov 18];48. Available from: https://www.thelancet.com/journals/lanepe/article/PIIS2666-7762(24)00335-1/fulltext

35. Chen Z, Liu X an, Kenny PJ. Central and peripheral actions of nicotine that influence blood glucose homeostasis and the development of diabetes. Pharmacol Res. 2023 Aug 1;194:106860.

36. Lin CC, Li CI, Liu CS, Lin CH, Yang SY, Li TC. Relationship between tobacco smoking and metabolic syndrome: a Mendelian randomisation analysis. BMC Endocr Disord. 2025 Mar 28;25(1):87.

37. Patel A, Figueredo VM. Alcohol and Cardiovascular Disease: Helpful or Hurtful. Rev Cardiovasc Med. 2023 Apr 19;24(4):121.

38. Ferriani LO, Silva DA, Viana MC. Atypical depression is associated with metabolic syndrome: a systematic review. Actas Esp Psiquiatr. 2022 Nov 1;50(6):266–75.

39. Shell AL, Crawford CA, Cyders MA, Hirsh AT, Stewart JC. Depressive disorder subtypes, depressive symptom clusters, and risk of obesity and diabetes: A systematic review. J Affect Disord. 2024 May 15;353:70–89.

40. Bazinet RP, Layé S. Polyunsaturated fatty acids and their metabolites in brain function and disease. Nat Rev Neurosci. 2014 Dec;15(12):771–85.

41. Walton NL, Antonoudiou P, Maguire JL. Neurosteroid influence on affective tone. Neurosci Biobehav Rev. 2023 Sep;152:105327.

42. Tao Z, Cheng Z. Hormonal regulation of metabolism-recent lessons learned from insulin and estrogen. Clin Sci Lond Engl 1979. 2023 Mar;137(6):415–34.

43. Sublette ME, Daray FM, Ganança L, Shaikh SR. The role of polyunsaturated fatty acids in the neurobiology of major depressive disorder and suicide risk. Mol Psychiatry. 2024 Feb;29(2):269–86.

