## Supplementary figures for "Clustering of major depressive disorder genetic instruments identifies distinct and directionally opposing effects on cardiometabolic risk"

Supplementary figure 1. MR-Clust results after applying Steiger filtering. Effect sizes are the estimates reported in the relevant GWASs. Error bars represent standard errors.

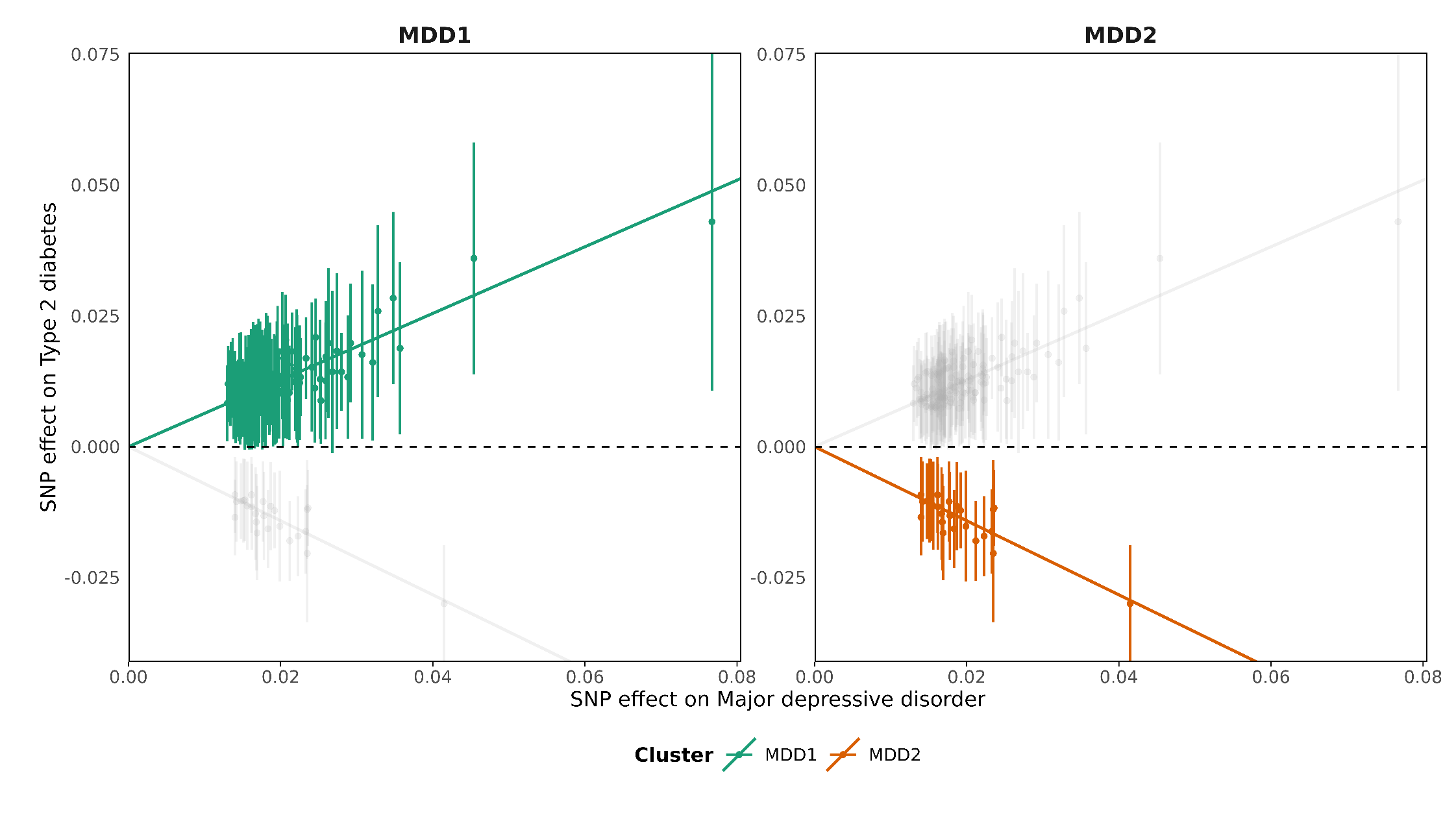

Supplementary Figure 2. Validation for the MDD-T2D SNP clusters using Two sample Mendelian randomisation with other T2D GWASs. Effect size represents Odds ratio per doubling of exposure genetic liability. Error bars represent 95% confidence intervals of the estimate. *, statistically significant after false discovery rate multiple testing correction.

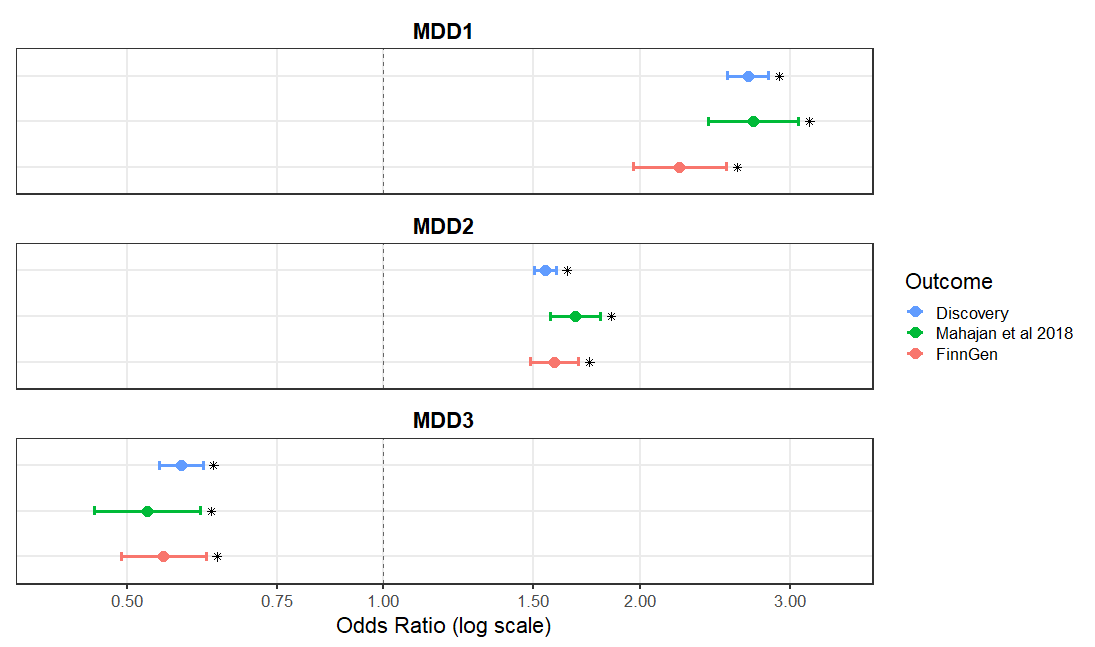

Supplementary Figure 3. Differential gene expression across 54 GTEx tissue types according to FUMA for the MDD1 SNP cluster.

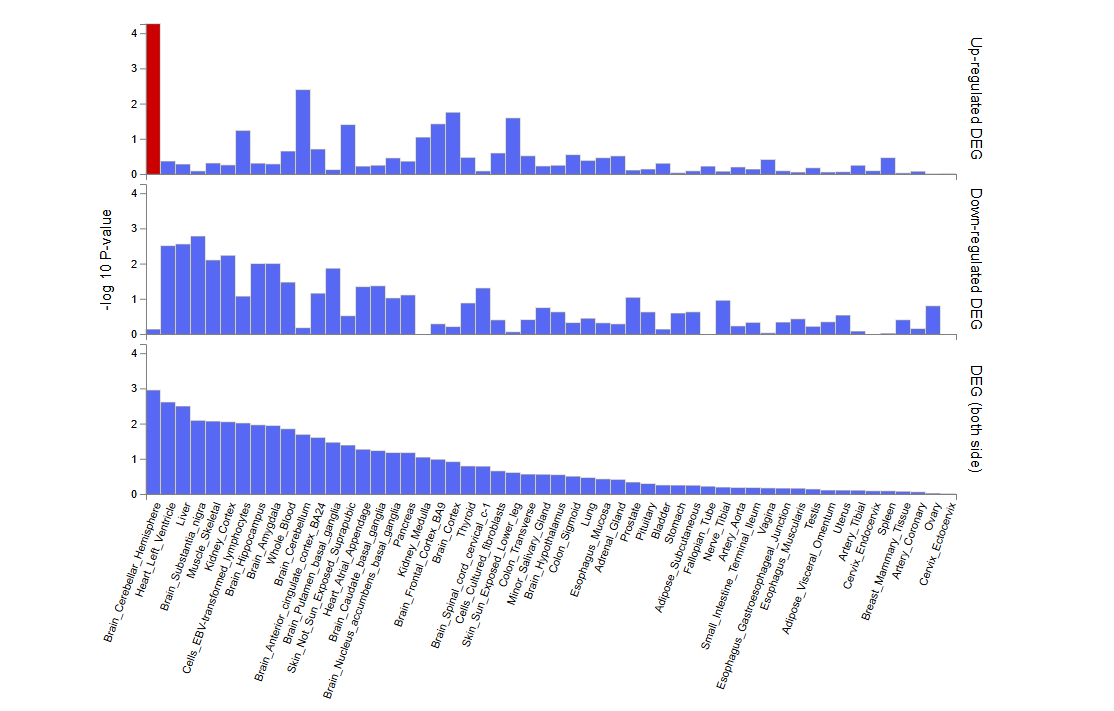

Supplementary Figure 4. Different	ial gene expression across 54 GTEx tissue types according to FUMA for the MDD1 SNP cluster.

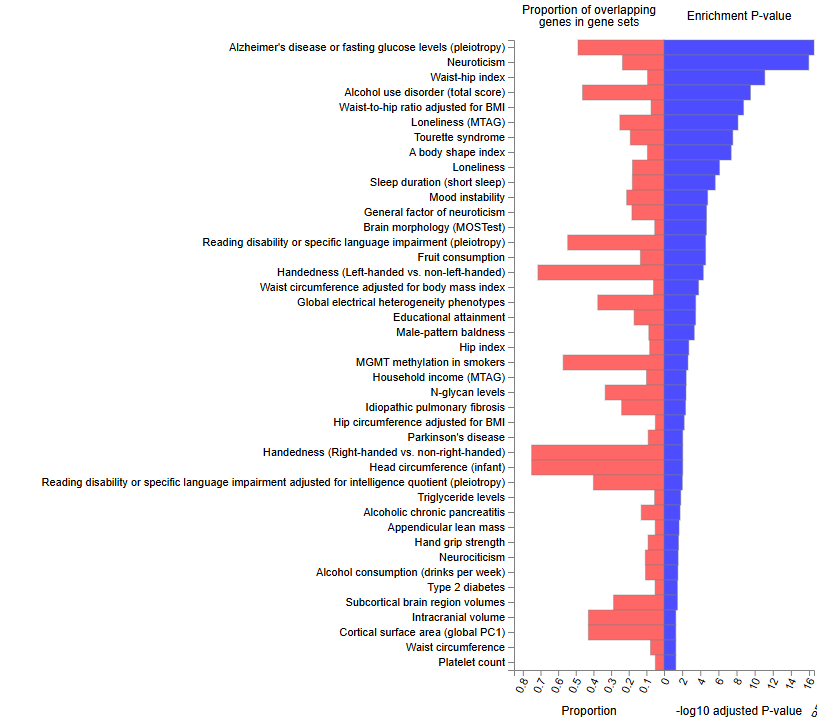

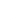

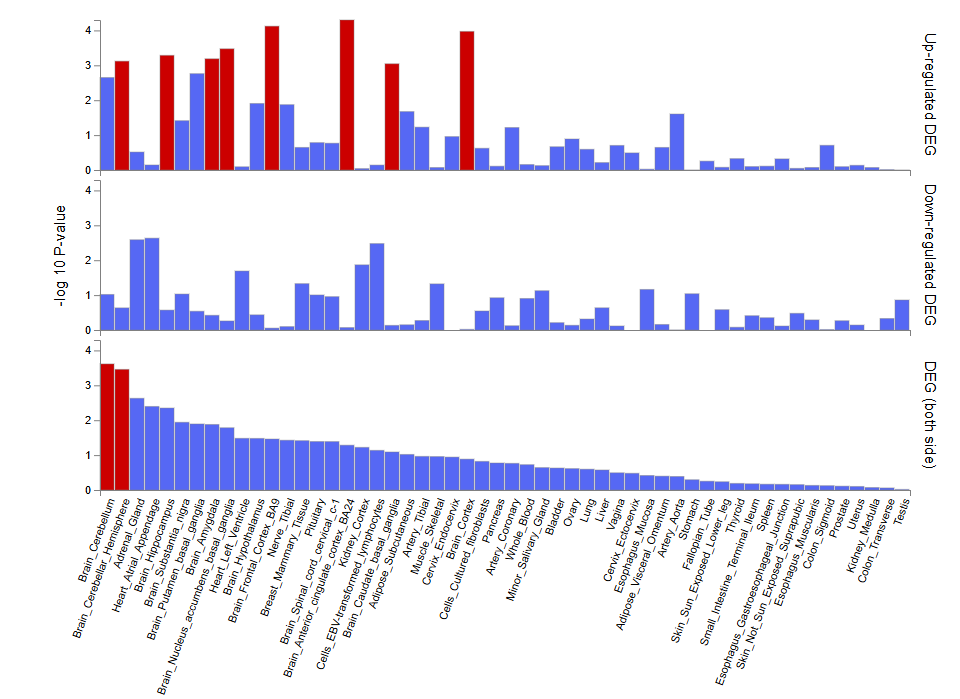
Supplementary Figure 5. Differential gene expression across 54 GTEx tissue types according to FUMA for the MDD2 SNP cluster.

Supplementary Figure 6. Differential gene expression across 54 GTEx tissue types according to FUMA for the MDD2 SNP cluster.

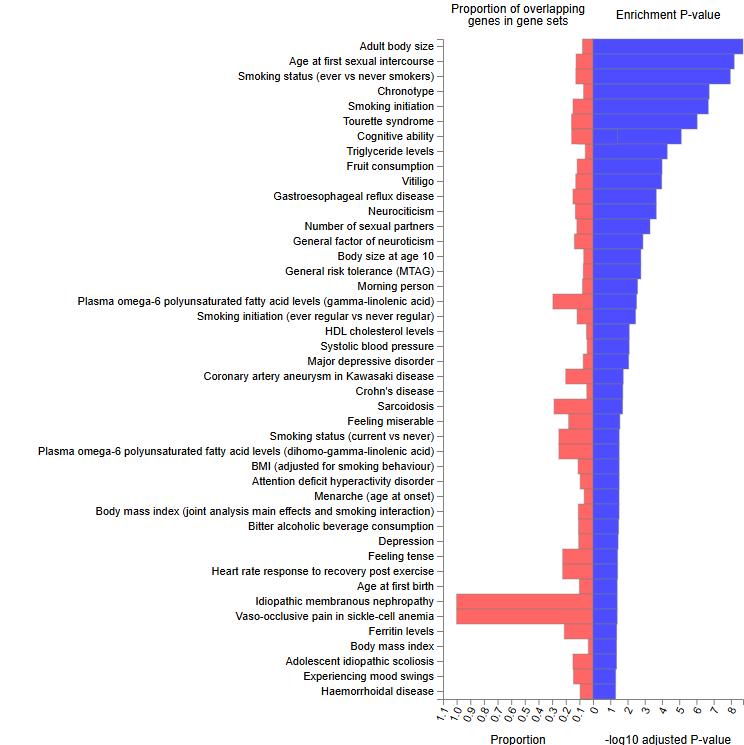

Supplementary Figure 7. Differential gene expression across 54 GTEx tissue types according to FUMA for the MDD3 SNP cluster.

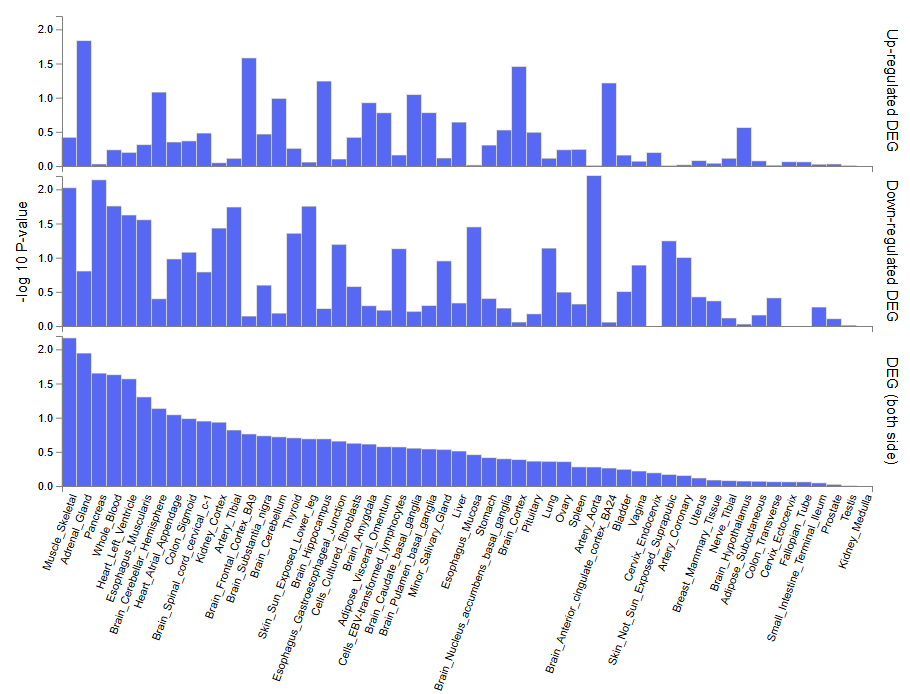

Supplementary Figure 8. Differential gene expression across 54 GTEx tissue types according to FUMA for the MDD3 SNP cluster.

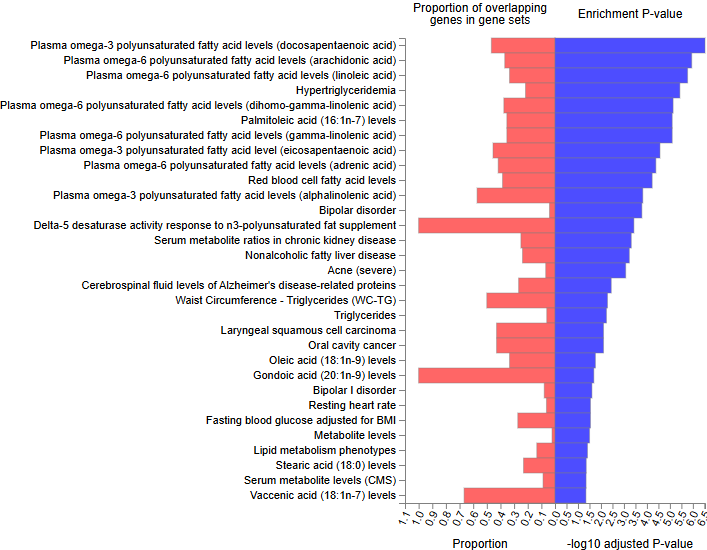

Supplementary figure 9. Pathway enrichment analysis for MDD3. A) reactome and B) Gene Ontology biological processes

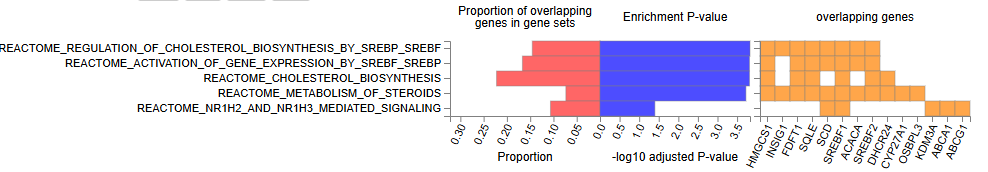

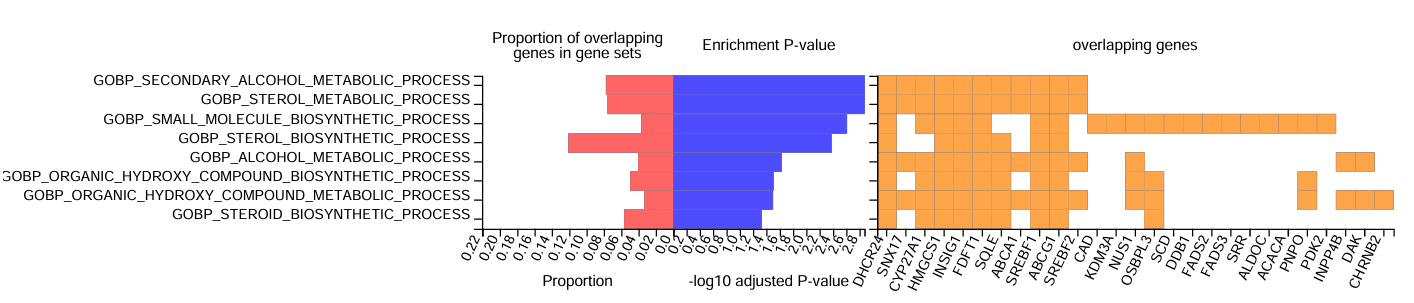

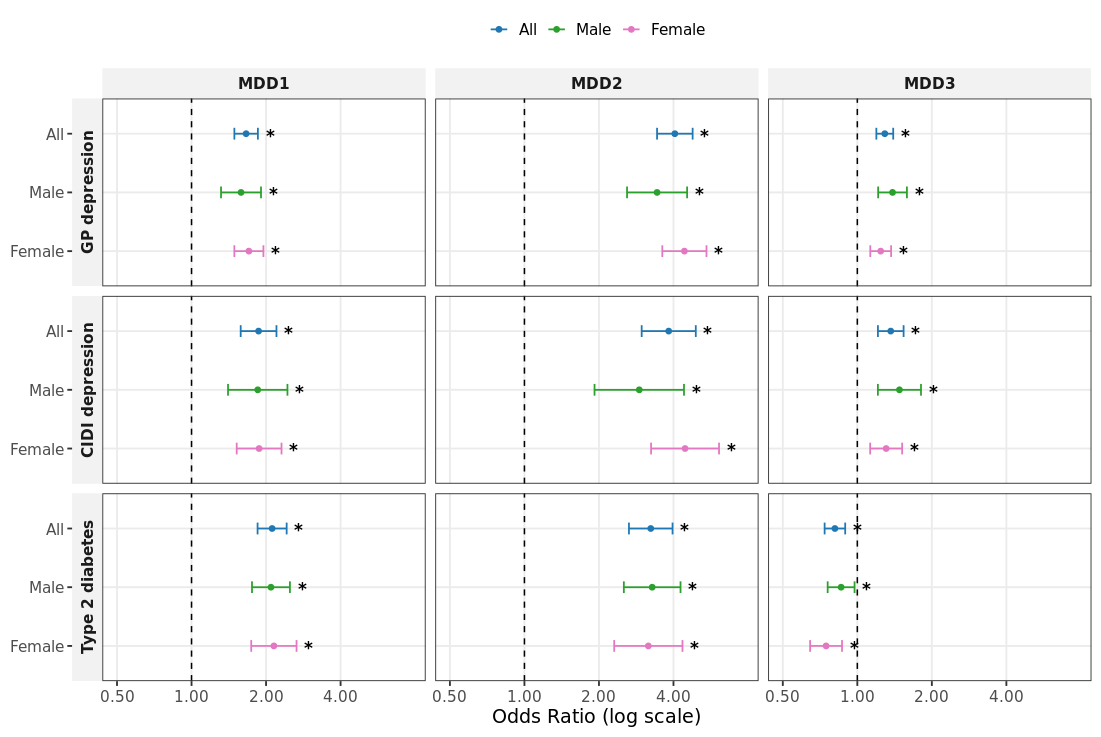
Supplementary Figure 10. Sex-stratified MDD-T2D SNP clusters associations with risk of GP-defined major depressive disorder, CIDI-SF defined MDD, and type 2 diabetes status. Effect sizes represent odds ratio per doubling in MDD genetic liability. Error bars represent 95% confidence intervals of the estimate. *, statistically significant after Benjamini-Hochberg false discovery rate adjustment (p < 0.05).

Supplementary Figure 11. Sex-stratified MDD-T2D SNP clusters associations with risk of different depression subtypes. Effect sizes represent odds ratio per doubling in MDD genetic liability. Error bars represent 95% confidence intervals of the estimate. *, statistically significant after Benjamini-Hochberg false discovery rate adjustment (p < 0.05). Sev dep, ever severely depressed; TR dep, treatment resistant depression; HWS dep, High WS depression; LWS, Low WS depression.

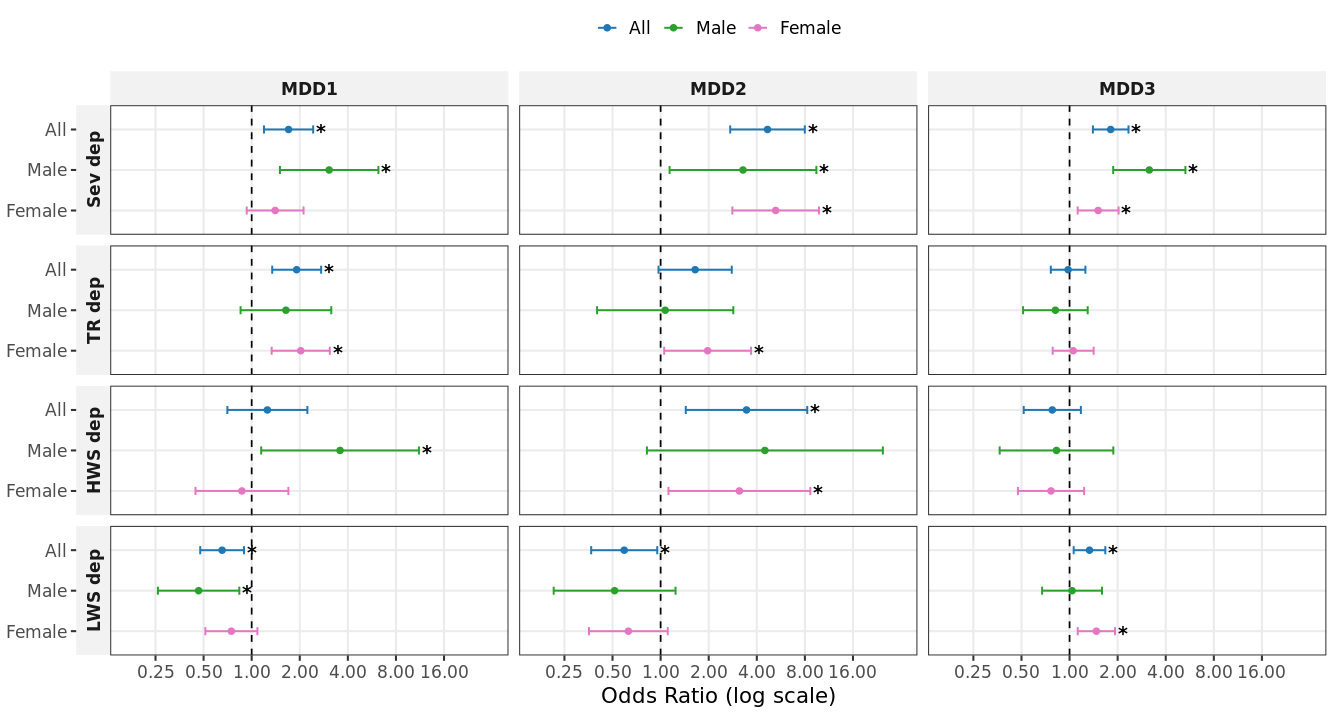

Supplementary figure 12. Sex-stratified causal effects of MDD-T2D SNP clusters on depression-related T2D risk factors. For easier comparison, the effect sizes are presented as Z scores standardised for the number of variants in each cluster. *, statistically significant after Benjamini-Hochberg FDR adjustment (p < 0.05). AUD, alcohol use disorder; Cigs, cigarettes; Mod-vig PA, Moderate to vigorous physical activity time; Light PA, light physical activity time; Sedentary, sedentary time; Education, education years; Deprivation, Townsend Deprivation Index.

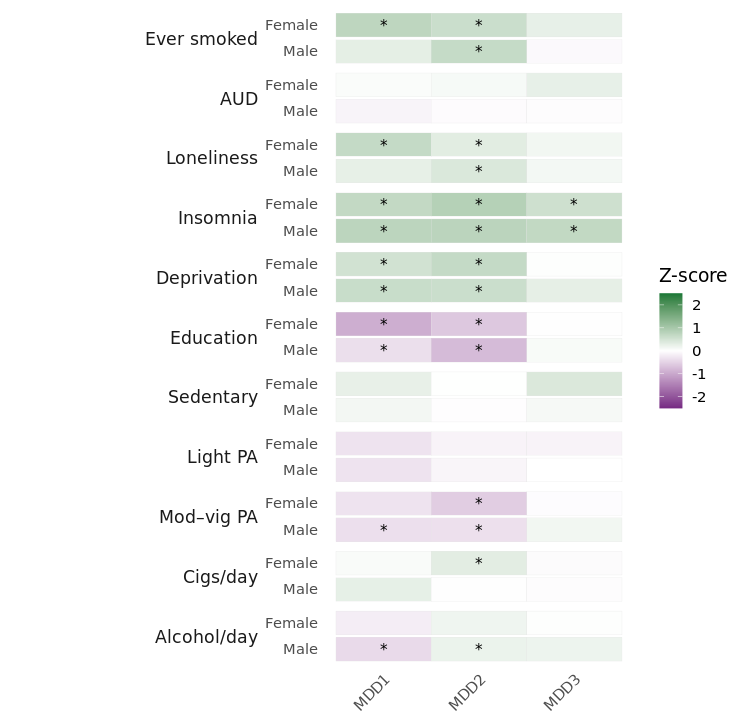

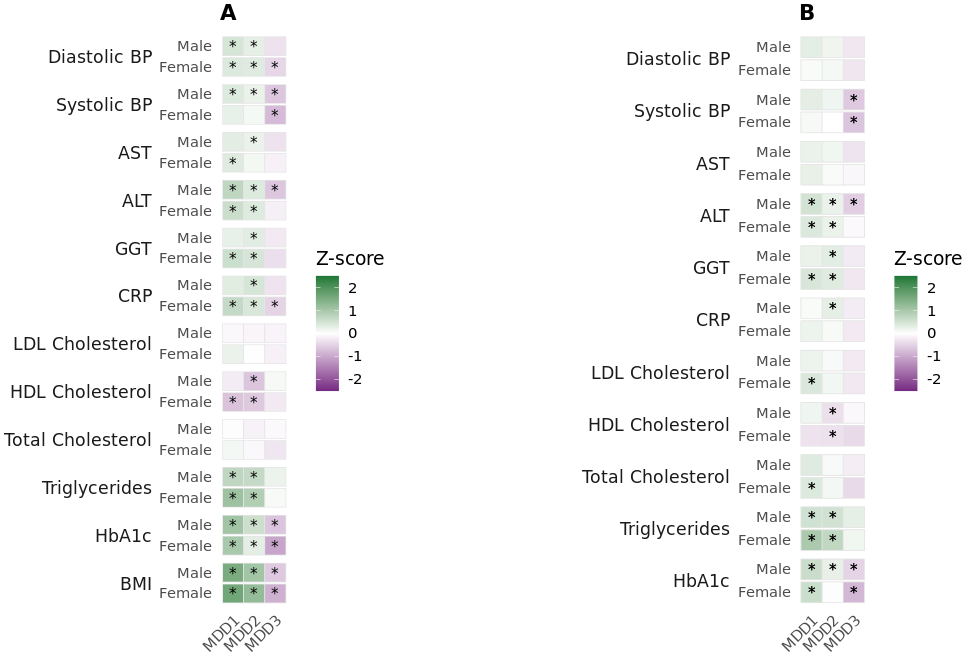
Supplementary Figure 13. Sex-stratified causal effects of MDD-T2D SNP clusters on cardiometabolic biomarkers A) unadjusted for BMI and B) adjusted for BMI. For easier comparison, the effect sizes are presented as Z scores standardised for the number of variants in each cluster. *, statistically significant after Benjamini-Hochberg FDR adjustment (p < 0.05). BMI, body mass index; HbA1c, glycated haemoglobin; CRP, C-reactive protein; GGT, γ-glutamyl transferase; ALT, alanine aminotransferase; AST, aspartate transaminase; BP, blood pressure.

Supplementary figure 14. Sex-stratified causal effects of MDD-T2D SNP clusters on different adiposity measures A) unadjusted for BMI and B) adjusted for BMI. For easier comparison, the effect sizes are presented as Z scores standardised for the number of variants in each cluster. *, statistically significant after Benjamini-Hochberg FDR adjustment (p < 0.05). Waist, waist circumference; hip, hip circumference; ASAT, abdominal subcutaneous adipose tissue; VAT, visceral adipose tissue; liver fat, percentage liver proton density fat fraction; pancreas fat, percentage pancreas proton density fat fraction; muscle fat, muscle fat infiltration; vol, volume.

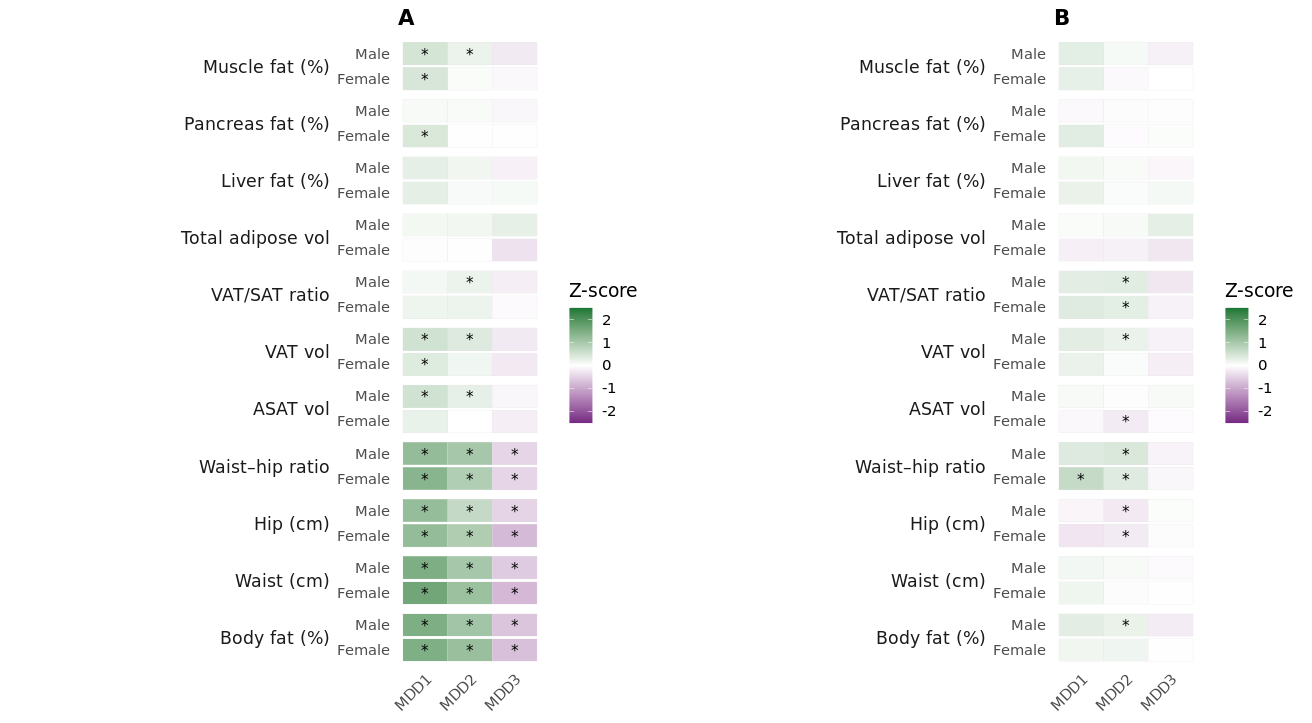
