## Supplementary methods for "Clustering of major depressive disorder genetic instruments identifies distinct and directionally opposing effects on cardiometabolic risk"

**Phenotype QC**

For all analyses, this work used a complete case analysis approach. Individuals that were missing either relevant covariates, exposure, or outcome data were excluded from analysis for each analysis respectively.

**Cluster validation phenotypes and depression subphenotypes**

*Type 2 diabetes*

Type 2 diabetes was derived from multiple sources within the UK Biobank, using data field 130709. Individuals were excluded as a type 2 diabetes case if they were prescribed insulin within the first year of diagnosis. In unrelated Europeans, there were 32,595 cases and 346,788 controls.

*GP derived MDD*

MDD within UKB was coded using the linked GP records. As per the Fabbri et al study (1) cases were defined if they had: a) at least two diagnostic codes for a depressive disorder, at any time point and b) no diagnostic code for bipolar disorder, psychotic disorders or substance abuse related disorders. In unrelated Europeans, we had 23,833 cases and 146,094 controls.

*Mental Health Questionnaire (MHQ) derived MDD and MDD subphenotypes*

A total of 145,668 participants completed the first UK Biobank Mental Health Questionnaire (MHQ). Depression phenotypes were derived using publicly available R code (<https://data.mendeley.com/datasets/kv677c2th4/3>), as previously described (2). Depression was assessed using the Composite International Diagnostic Interview Short Form (CIDI-SF) and the Patient Health Questionnaire-9 (PHQ-9). From the CIDI-SF, we generated a binary measure indicating lifetime major depression and a continuous measure reflecting the severity of lifetime depression. Ever being “severely depressed” was defined as having a total CIDI-SF score of 8, and reporting “a lot” of impairment on the CIDI-SF impairment question. For depressed ever MDD phenotype, we had 29563 cases and 70081 controls. In addition, we also used the MHQ data to derive the Low weight and sleep (low WS) and high weight and sleep (high WS) phenotypes, as described previously (3).

*Treatment resistant depression (TRD)*

Treatment-resistant depression (TRD) was defined using linked GP electronic health records and prescription data, following the approach described by Fabbri et al. (1). These records provide detailed information on antidepressant prescriptions. TRD was coded when an individual had been prescribed at least two different antidepressants, each for a minimum duration of six weeks. This 6-week threshold, which is more conservative than the four-week change has been recommended in prescribing guidelines, was chosen to reduce the likelihood that medication changes were due to side effects, while still allowing sufficient time to assess treatment efficacy. In unrelated Europeans we had 2,216 cases, 22,748 controls.

**Depression-related cardiometabolic risk factors**

*Insomnia*

Insomnia was defined using UK Biobank baseline data field 1200, which has been validated as a reliable proxy for insomnia disorder (4). Participants were asked: “Do you have trouble falling asleep at night or do you wake up in the middle of the night?” with four possible responses: ‘never/rarely’, ‘sometimes’, ‘usually’, and ‘prefer not to answer’. Those selecting ‘prefer not to answer’ were excluded. For analysis, the variable was dichotomized, with individuals responding ‘usually’ classified as having insomnia. Individuals who answered ‘sometimes’ and “never/rarely’ were considered controls or no insomnia. In unrelated European, we had 271,897 control individuals and 107,256 insomnia cases.

*Loneliness and alcohol use disorder*

Loneliness and alcohol use disorder were derived using publicly available R code (<https://data.mendeley.com/datasets/kv677c2th4/3>), as previously described (2).

*Smoking status*

1. **Cigarettes per day:** Cigarettes per day was derived from various UKB smoking variables. If someone’s smoking status (data field 20116) is 0, 1, and 2, they are considered as “never smoker”, “current smokers” and “past smokers” respectively. For current smoker, their number of cigarettes per day count was then taken from the self-reported number of cigarettes currently smoked per day (data field 3456). For past smokers, it was taken from the self-reported number of cigarettes previously smoked per day (data field 2887). Never smokers were assigned a value of zero. We had 27599 people for this variable.
2. **Ever smoked**: This variable was derived from UKB data field 20160. We had total

378168 individuals under this category.

*Alcohol use (Units per day)*

Units per day were derived from UK Biobank self-reported alcohol intake. For weekly drinkers {data fields 1568 (red wine), 1578 (white wine), 1588 (beer/cider), 1598 (spirits), 1608 (fortified wine)}, weekly units were calculated using following standard UK conversion factors:

Weekly units=1.5 × (red wine + white wine) + 2.8 × beer/cider + spirits + fortified wine

Each additional alcoholic drinks (data field 5364) was then added at 1.5 units each, then divided by 7 to obtain units per day.

For monthly drinkers {data field 4407(red wine), 4418 (white wine), 4429 (beer/cider), 4440 (spirits), 4451 (fortified wine), 4462 (additional alcoholic drinks)}, total monthly units were calculated similarly as explained above and divided by 30.4 to get units per day metrics. Participants reporting never drinking (data field 15586) were assigned 0 units/day. We had 320,066 individuals in this category.

*Accelerometry-defined physical activity*

These metrics were derived from the actigraphy devices (Axivity AX3) which was worn by 103,711 individuals from the UK Biobank for up to 7 days. We had 81405 individuals available for this variable and inverse normalised physical activity and sedentary time metrics was used for downstream analysis.

*Educational attainment*

We used years of education as a proxy of education, which is an inferred length of total education duration derived using multiple UK Biobank fields, as previously described (5).

*Socio-economic status*

Townsend deprivation index was used as a proxy for socio-economic status, which was collected at UK Biobank recruitment in data field 22189. All individuals are assigned a score based on their post code at time of recruitment.

**Cardiometabolic biomarkers**

*HbA1c*

Glycated Haemoglobin (HbA1c) values were derived using data field 30750. These values were not corrected for anti-diabetic medication use.

*Body mass index*

Body mass index values were derived using field 21022, which are generated using participant weight and height at recruitment. Individuals without either a height or weight measurements are omitted.

*Blood lipid levels*

In total, we considered four lipid levels for analysis: High density lipoprotein cholesterol (HDL; field 23406), low density lipoprotein (LDL; field 23405), total triglycerides (field 23407), and total cholesterol (field 23400).

We have also accounted for statins (and other lipid-lowering medication) while curating the lipid biomarker data in the following way:

Table showing adjustment applied to the lipid biomarkers for individuals in UKB on lipid lowering medication.

|  | TC | LDL | HDL | Trig |  |
| --- | --- | --- | --- | --- | --- |
| Statins | 1.2687 | 1.4068 | 0.9655 | 1.1563 | Based on -Cholesterol Treatment Trialists’ (CTT) Collaboration. Efficacy and safety of more intensive lowering of LDL cholesterol: a meta-analysis of data from 170 000 participants in 26 randomised trials. Lancet 2010; published online November 9, 2010. DOI:10.1016/S0140-6736(10)61350-5 |
| Ezetimibe | 1.135 | 1.186 | 0.970 | 1.086 | based on - Ezetimibe monotherapy for cholesterol lowering in 2,722 people: systematic review and meta‐analysis of randomized controlled trials |

*Liver biomarkers*

We used three commonly measured liver enzyme biomarkers as a proxy of liver function: Alanine aminotransferase (field 30620), Aspartate aminotransferase (field 30650), and γ-glutamyl transferase (field 30730).

*Blood pressure*

We considered both systolic blood pressure (field 4080) and diastolic blood pressure (field 4079) for analysis. These values were adjusted for blood pressure lowering medication use according to previous studies (6), where systolic blood pressure was increased by 10 mmHg, and diastolic blood pressure was increased by 5mmHg.

*Systemic inflammation*

We used C-reactive protein as a global biomarker proxy for systemic inflammation (field 30710)

**Adiposity traits**

*Body fat percentage*

Body fat percentage determined using bioelectrical impedance was used as a measure of overall adiposity (field 23099).

*Waist-hip ratio*

Waist-hip ratio was used as a measure of centralised adiposity and was calculated by diving the waist circumference (field 48) by the hip circumference (field 49). Individuals without both measurements at a single data collection time point were excluded.

*Visceral and abdominal subcutaneous tissue volume and VAT:aSAT ratio*

Visceral (VAT; 55,955) and abdominal subcutaneous tissue volume (aSAT; field 22408) were measured using abdominal magnetic resonance imagery. VAT:aSAT ratio was calculated by dividing the VAT tissue volume by the aSAT volume. In total, 55,458 individuals with both measurements were included for analyses.

*Total adipose tissue volume*

Total adipose tissue (field 22415) is the total adipose tissue, as measured by Magnetic resonance imagery, between the bottom of the thigh muscles to the top of the vertebrae T9. In total, 6,544 individuals were included.

*Hepatic and pancreatic adipose tissue deposition*

Liver (field 21088) and pancreas (field 21090) adipose tissue deposition were determined by magnetic resonance imagery.

*Thigh muscle fat infiltration*

Muscle fat infiltration (field 22435) was defined as the fat fraction in viable anterior thigh muscle tissue.
